# Social predictors of food insecurity during the stay-at-home order due to the COVID-19 pandemic in Peru. Results from a cross-sectional web-based survey

**DOI:** 10.1101/2021.02.06.21251221

**Authors:** Jorge L. Cañari-Casaño, Omaira Cochachin-Henostroza, Oliver A. Elorreaga, Gandy Dolores-Maldonado, Anthony Aquino-Ramírez, Sindy Huaman-Gil, Juan P. Giribaldi-Sierralta, Juan Pablo Aparco, Daniel A. Antiporta, Mary E. Penny

## Abstract

**Background:** Stay-at-home orders and social distancing have been implemented as the primary tools to reduce the spread of severe acute respiratory syndrome coronavirus 2 (SARS-CoV-2). However, this approach has indirectly lead to the unemployment of 2·3 million Peruvians, in Lima, Perú alone. As a result, the risk of food insecurity may have increased, especially in low-income families who rely on a daily wage. This study estimates the prevalence of moderate or severe food insecurity (MSFI) and identifies the associated factors that explain this outcome during the stay-at-home order.

**Methods:** A cross-sectional web-based survey, with non-probabilistic sampling, was conducted between May 18 and June 30, 2020, during the stay-at-home order in Peru. We used social media advertisements on Facebook to reach 18-59-year-olds living in Peru. MSFI was assessed using the Food Insecurity Experience Scale (FIES). Rasch model methodology requirements were considered, and factors associated with MSFI were selected using stepwise forward selection. A Poisson generalized linear model (Poisson GLM), with log link function, was employed to estimate adjusted prevalence ratios (aPR).

**Findings:** This analysis is based on 1846 replies. The prevalence of MSFI was 23·2%, and FIES proved to be an acceptable instrument with reliability 0·72 and infit 0·8-1·3. People more likely to experience MSFI were those with low income (less than 255 US$/month) in the pre-pandemic period (aPR 3·77; 95%CI, 1·98-7·16), those whose income was significantly reduced during the pandemic period (aPR 2·27; 95%CI, 1·55-3·31), and those whose savings ran out in less than 21 days (aPR 1·86; 95%CI, 1·43-2·42). Likewise, heads of households (aPR 1·20; 95%CI, 1·00-1·44) and those with probable SARS-CoV2 cases as relatives (aPR 1·29; 95%CI, 1·05-1·58) were at an increased risk of MSFI. Additionally, those who perceived losing weight during the pandemic (aPR 1·21; 95%CI, 1·01-1·45), and increases in processed foods prices (aPR 1·31; 95%CI, 1·08-1·59), and eating less minimally processed food (aPR 1·82; 95%CI, 1·48-2·24) were more likely to experience MSFI.

**Interpretation:** People most at risk of MSFI were those in a critical economic situation before and during the pandemic. Social protection policies should be reinforced to prevent or mitigate these adverse effects.

**Funding:** None.

## INTRODUCTION

By the end of January 2020, the world had experienced over 100 million SARS-CoV2 cases, of whom approximately 2.3 million have died.^(1)^ Peru has had around 1 million cases and more than 41,000 have died from SARS-CoV2.^(2)^ Moreover, Peru is one of the countries with the highest SARS-CoV2 mortality rate in the world.^(3)^ After declaring a global health emergency,^(4)^ the World Health Organization (WHO) recommended social distancing and closing public and private services, among other measures to contain the spread of SARS-CoV2.

Two principal measures adopted by many countries were stay-at-home orders and border closures (i.e., national lockdowns). In Peru, these were implemented from March 16 to June 30, 2020. People were only allowed to go out for essential activities, for instance to buy food and medicine. However, due to the precarious labour market in Peru, where informal labour constitutes 50% of all labour,^(5)^ and pre-pandemic poverty the purchasing power of Peruvian families was severely affected by the lockdown. The Peruvian government tried to help this vulnerable population by giving social aid (i.e., financial stimulus to families, early withdrawal from the private retirement pension, and food donations) to face the stay-at-home orders’ side effects. It has been suggested that this was not enough due to inefficient distribution and the already critical situation exacerbated by COVID-19 pandemic.^(6)^

stay-at-home orders could affect food insecurity due to the resulting economic crisis, massive loss of jobs, high food prices, and growing demand for medical care.^(7)^ These results may generate short- and medium-term adverse health effects.^(7)^ Recent research on the short-term impacts of stay-at-home orders supports this idea. One of the first studies in Peru reported a considerable income reduction after stay-at-home orders in 37% of the participants,^(8)^ and over 2·3 million lost jobs in Lima alone. ^(9)^ Similar occurrences have been reported outside Peru.^(11-10)^ Furthermore, higher food prices, changes in diets, increased depression, anxiety and physical violence against women have all been associated with stay-at-home orders. ^(11,12,13)^

Recent studies have also reported an increase in food insecurity after implementation of stay-at-home orders,^(14-16)^ which worsens as the crisis intensifies.^(11)^ These results are alarming, especially considering that the World Food Program estimated the number of people experiencing food insecurity will increase to 265 million by the end of 2020,^(17)^ accompanied by potential negative effects on health and household relations.^(7)^

While MSFI was estimated at 29.9% between 2014-2016 in Peru,^(18)^ little data exist regarding how stay-at-orders have affected MSFI. The present study aimed to estimate the prevalence of MSFI and determine the factors associated with MSFI in our study population.

#### Research in context

##### Evidence before this study

Stay-at-home orders established to stop the spread of the COVID-19 pandemic have caused profound adverse effects in low- and middle-income countries such as Peru, specifically in economically vulnerable populations, who are most at risk for food insecurity. We searched PubMed servers up to Jan 20, 2020, for peer-reviewed articles published using the terms “(food insecurity OR food security) AND (COVID-19 OR SARS-CoV2) OR (stay-at-home order OR Quarantine OR lockdown)”, without data or language restrictions. Our search found few studies that have examined the direct or indirect impact of stay-at-home orders on moderate-severe food insecurity (MSFI). However, the available evidence is scarce in low- and middle-income settings, especially Latin American countries such as Peru.

##### Added value of this study

To our knowledge, this study provides one of the first pieces of evidence about the prevalence of MSFI, measured using the Food Insecurity Experience Scale (FIES), and its associated factors during the stay-at-home order in Peru. Our findings agree with the growing scientific literature reporting that those most at risk of MSFI occupy tenuous economic positions before and during the pandemic.

##### Implications of all the available evidence

It is urgent to implement social protection policies that aim to strengthen real-time systems focusing on identifying and providing social assistance to vulnerable populations most at risk of MSFI. These systems should consider that non-poor households can quickly descend into poverty due to the inability of day laborers to work during stay-at-home orders. It is also essential to identify appropriate social aid strategies (money transfers, food donations, communal pots of food, etc.) according to the vulnerable population’s characteristics and state of the pandemic. These recommendations may mitigate the adverse effects of stay-at-home orders on food MSFI.

## METHODS

### Study design and participants

In this cross-sectional study an online survey administered through the Qualtrics platform was used to estimate the prevalence of and factors associated with MSFI. The survey was administered to Peruvian adults, 18 to 59 years old who had internet and social media (Facebook) access by mobile devices or computers.

### Procedures

The survey was accessible between May 18 and June 30 during the stay-at-home order. The dissemination of this study was done by: (1) paid advertisements from the “Colegio de Nutricionistas del Perú” official social media site; (2) social media posts by public and private universities and Peruvian research centres, and (3) by dissemination of the online survey link by participants through their personal social media profile or through their WhatsApp groups and contacts.

Participants who clicked on the link were redirected to a Qualtrics survey platform to read more about the study and consent before starting the survey. The survey could be answered more than once, and if this happened, participants had to mention why they were repeating the survey. Participants who completed the study were invited to download educational material on “Healthy Eating” and asked to share the survey link through their social media and WhatsApp contacts.

The online survey collected sociodemographic information (i.e., place of residence, sex, age, marital status, nationality, educational level, employment status, whether they suffered from a chronic disease or disability, SARS-CoV2 symptoms or any relative that tested positive for SARS-CoV2 in participant environment); FIES; household information (i.e., number of household members, average monthly income, economic condition during stay-at-home order, emergency savings, government financial support, water connection and drainage); perception about food prices (i.e., prices relative to pre-pandemic period: “lower price”, “same price”, “higher price”, “do not know/prefer not to answer”); perception about food consumption (i.e., “eat less”, “eat the same”, “eat more”, “do not eat/prefer not to answer”); perception about places to buy and waiting time, and perception about body weight status before and during the pandemic using body shape images were included in the questionnaire. To validate the survey questionnaire, a pilot study was conducted by phone call to 23 people of different age groups.

### Food Insecurity Measurement

We used the Food Insecurity Experience Scale (FIES) at the individual level to determine the prevalence of MSFI. The FIES is an eight-item questionnaire with dichotomous responses (i.e., Yes or No) about experiences with food insecurity at the household or individual level.

The FIES was developed by Food and Agriculture Organization’s (FAO) Voices of the Hungry (VoH) project and was validated in 151 countries with Gallup World Poll (GWP) data in 2014.^(19-21)^ Several studies have used FIES to determine food insecurity in different fields and context.^(22-28)^

We adapted the FIES questionnaire to the Peruvian context taking into consideration a study by Vargas and Penny,^(29)^ and a pilot study (n=23) where content and understanding were evaluated. Minor modifications were done to the formulation of the questions and the whole questionnaire was translated into Spanish (Table 1).

**Table 1:** Food Insecurity Experiences Scale (FIES)^1^.

| Order | Questions | Items | Possible answers |
| --- | --- | --- | --- |
| Q1 | During the last month of quarantine, was there a time when, because of lack of money or other resources, you were worried you would not have enough food to eat? | WORRIED | 1= Yes<br>2= No<br>3= Don’t Know<br>4=Prefer not to answer |
| Q2 | During the last month of quarantine, was there a time when, because of lack of money or other resources, you could not eat healthy and nutritious food? For example: meat, fish, fruits, or vegetables. | HEALTHY | 1= Yes<br>2= No<br>3= Don’t Know<br>4=Prefer not to answer |
| Q3 | During the last month of quarantine, was there a time when, because of lack of money or other resources, you had to eat only a few kinds of foods? | FEWFOODS | 1= Yes<br>2= No<br>3= Don't Know<br>4=Prefer not to answer |
| Q4 | During the last month of quarantine, was there a time when, because of lack of money or other resources, you had to reduce the number of meals per day because you did not have enough food? For example, skip breakfast, lunch, or dinner? | SKIPPED | 1= Yes<br>2= No<br>3= Don't Know<br>4=Prefer not to answer |
| Q5 | During the last month of quarantine, was there a time when, because of lack of money or other resources, you had to eat less than you thought you should eat? | ATELESS | 1= Yes<br>2= No<br>3= Don't Know<br>4=Prefer not to answer |
| Q6 | During the last month of quarantine, was there a time when, because of lack of money or other resources, your food ran out in your household? | RANOUT | 1= Yes<br>2= No<br>3= Don't Know<br>4=Prefer not to answer |
| Q7 | During the last month of quarantine, was there a time when, because of lack of money or other resources, you went to sleep hungry because there was not enough food? | HUNGRY | 1= Yes<br>2= No<br>3= Don't Know<br>4=Prefer not to answer |
| Q8 | During the last month of quarantine, was there a time when, because of lack of money or other resources, you went without eating for a whole day? | WHLDAY | 1= Yes<br>2= No<br>3= Don't Know<br>4=Prefer not to answer |

<sup>1</sup> The Spanish version of the FIES is provided in the supplementary material.

To analyze and determine MSFI, we only considered people who responded positively (value: 1) or negatively (value: 0) to all questions on the FIES. Since the FIES scale fulfilled all Rasch model assumptions (reliability and infit), a raw score was obtained (i.e., sum of affirmative responses). Individuals with raw scores between 4-8 were classified as MSFI and those with scores of 0-3 were classified as not having MSFI.^(30)^ Similar cut off points were used to determine MSFI in other studies ^(22, 26, 31, 32)^

### Covariates

Sociodemographic variables including age, sex, place of residence, educational level, if respondent was a head of household, and number of household members were considered. Household income level before and during the pandemic were related to an individual’s financial status and use and duration of savings during the stay-at-home order. Variables related to social assistance included support given by the government or any institution. Variables used to understand changes since the emergency began, included changes in location where food was bought and principal food groups consumed (i.e., healthy or minimally processed food, culinary ingredients, processed food, and ultra-processed foods). Body weight perceptions before and during the pandemic were evaluated using the Stunkard scale.^(33)^ Finally, variables related to COVID-19 pandemic were also considered; specifically, participants reporting a SARS-CoV2 diagnosis, loss of smell/taste, or whose household members had or had had SARS-CoV2.

### Data Analysis

Rash model assumptions for the FIES scale were evaluated using the RM.weight package in R statistical software,^(21, 34)^ where performance of an eight-item questionnaire was assessed utilizing infit statistic and the ability to distinguish different levels of food insecurity in participants was evaluated with the reliability of FIES scale. After verifying the fulfilment of the FIES scale’s assumptions, food insecurity was determined using the raw score (the sum of affirmative responses). Descriptive analysis of categorical variables and bivariate analysis between MSFI and other predictor variables were performed by proportions and chi-square test, respectively. Factors associated with the predictor variables and MSFI were determined using forward stepwise selection, starting with the most significant predictor variable. We used Poisson generalized linear models (Poisson GLMs) with log link function, to estimate adjusted prevalence ratios (aPR) fitted at the departmental level. Data analysis was performed using Stata 15·0 (Stata Corp., College Station, Texas – United States) and software program R studio (Version 4.0.4; R Foundation for Statistical Computing, Vienna, Austria).

### Ethical Issues

The study was approved by the human ethics committee of “Instituto de Investigación Nutricional” (CIEI-IIN), Lima – Peru, N° 394-2020/CIEI-IIN.

## RESULTS

A total of 2643 people answered the study’s informed consent, and 116 declined to participate in the study. Fifty-two participants were excluded because we could not determine if they had taken the survey more than once or they retook the survey for an inadequate reason and 629 participants because they did not answer all the questions on the FIES scale. The final sample considered for the analysis was 1846 participants (Figure 1).

**Figure 1:**
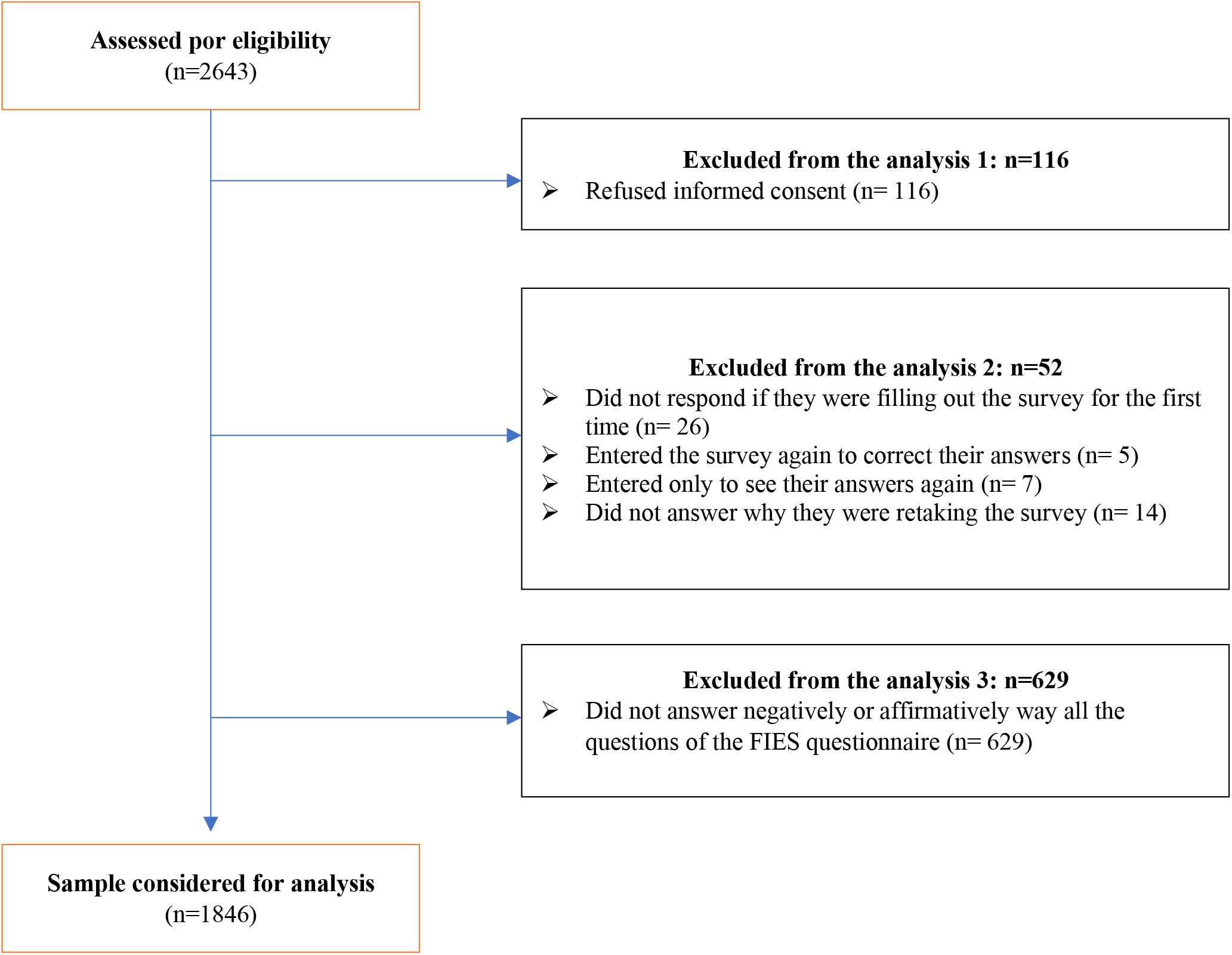
Flow diagram.

Table 2 shows the infit statistic and the Rasch model parameters. Infit values were acceptable, and Rasch model reliability in the FIES scale was 0·72 (data not shown in the table) which was also acceptable (greater than 0·7).

**Table 2:** Evaluation of the assumptions of the Rash model of the FIES scale.

| Item | Item_sev.* | St.err.† | Infit‡ | Outfit |
| --- | --- | --- | --- | --- |
| WORRIED | -2.37100181 | 0.0804195 | 1.2714576 | 2.2715038 |
| FEWFOOD | -1.99919744 | 0.0785325 | 0.8542389 | 0.7456257 |
| HEALTHY | -0.72783574 | 0.0818627 | 1.0229524 | 0.9484038 |
| ATELESS | -0.8382067 | 0.08112009 | 0.8300268 | 0.7194007 |
| SKIPPED | 0.02735437 | 0.0891645 | 0.9363935 | 0.8611874 |
| RUNOUT | 0.69432414 | 0.09915824 | 1.1374597 | 1.2012408 |
| HUNGRY | 1.31138247 | 0.1125294 | 0.8408805 | 0.7309963 |
| WHLDAY | 3.90325585 | 0.25963247 | 1.0598114 | 0.7132612 |

\*Item\_sev: Item severity. †St.err: Standard error. ‡Infit: Infit statistic. Acceptable values are in the range of 0.7 - 0.13.

Most of the participants were women (74·9%) with a technical or university education (85·3%). About a third were heads of household (34·2%) and lived-in provinces other than Metropolitan Lima (32·2%). Regarding economic factors, a low proportion of people (12·4%) received less than the minimum wage in Peru (less than 255 US$/month) before the pandemic and only 23·4% continued receiving income during the stay-at-home orders. On the other hand, over half of participants (56·7%) had emergency savings left to address the pandemic, while 16% and 9·5% of people ran out of savings in 22-35 days and less than 21 days since the start of the stay-at-home order, respectively.

As for factors related explicitly to the stay-at-home order, we found 17% of people had relatives with probable cases of SARS-CoV2, 30% stayed at home without working but had worked in the pre-pandemic period, 38·1% were working during the stay at home period, and 26·8% were not working before or during the stay at home order. 71·6% reported that the process to buy food took more time than before, and only 23·4% mentioned they received some support (include the government and private support) during the COVID-19 pandemic. In relation to body weight perceptions, 24·9% people perceived losing weight, 32·1% maintained their weight, and 30·7% gained weight. Regarding natural or minimally processed food consumption, 30% of participants mentioned that they ate less of this food group, and 39% ate more. It was found that 23·3% of participants experienced MSFI (Table 3).

**Table 3.**
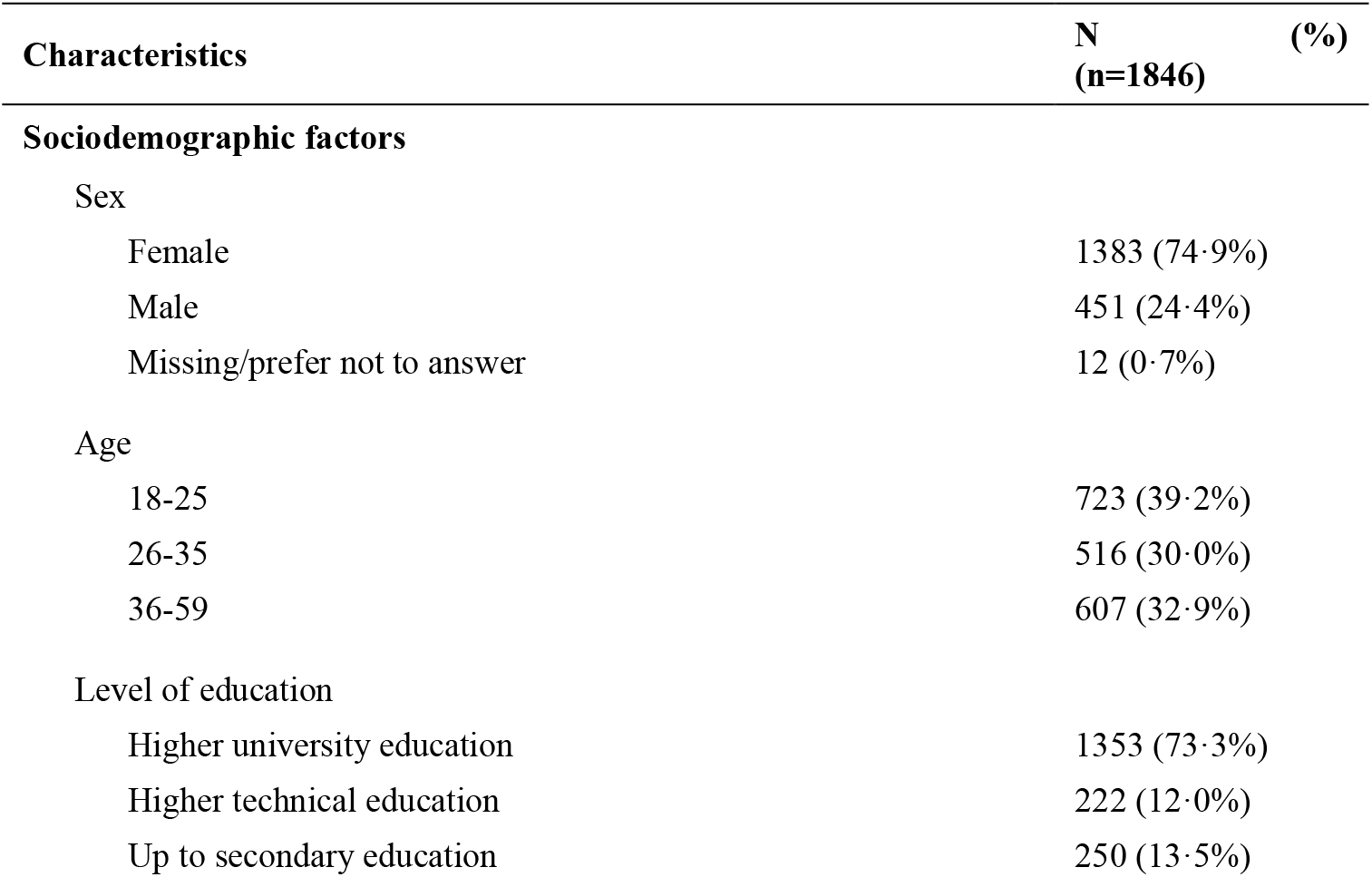
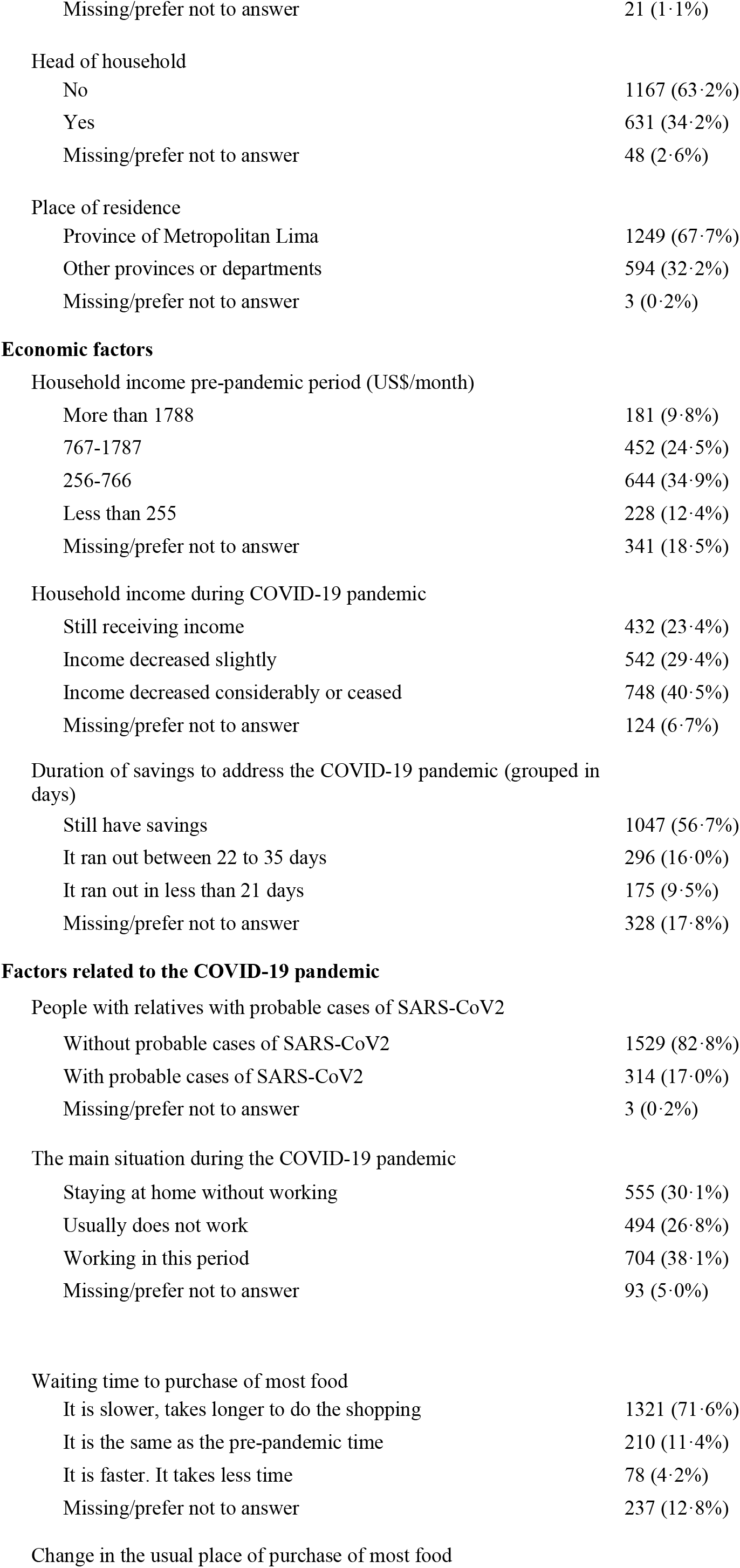
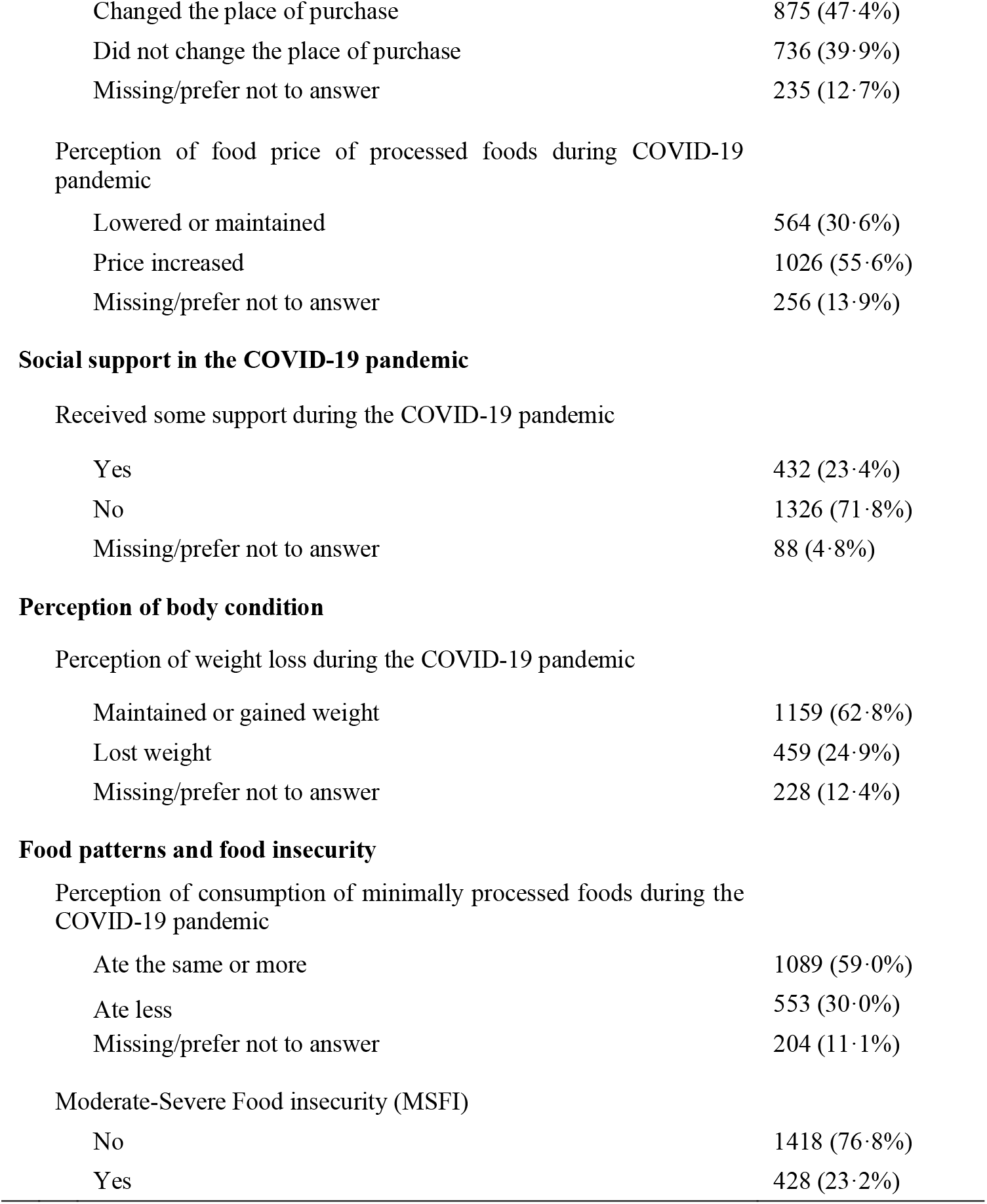
General characteristics of the population studied.

| Characteristics | N<br>(n=1846) | (%) |
| --- | --- | --- |
| **Sociodemographic factors** |  |  |
| Sex |  |  |
| Female | 1383 | (74.9%) |
| Male | 451 | (24.4%) |
| Missing/prefer not to answer | 12 | (0.7%) |
| Age |  |  |
| 18-25 | 723 | (39.2%) |
| 26-35 | 516 | (30.0%) |
| 36-59 | 607 | (32.9%) |
| Level of education |  |  |
| Higher university education | 1353 | (73.3%) |
| Higher technical education | 222 | (12.0%) |
| Up to secondary education | 250 | (13.5%) |

|  |  |
| --- | --- |
| Missing/prefer not to answer | 21 (1.1%) |
| Head of household |  |
| No | 1167 (63.2%) |
| Yes | 631 (34.2%) |
| Missing/prefer not to answer | 48 (2.6%) |
| Place of residence |  |
| Province of Metropolitan Lima | 1249 (67.7%) |
| Other provinces or departments | 594 (32.2%) |
| Missing/prefer not to answer | 3 (0.2%) |
| **Economic factors** |  |
| Household income pre-pandemic period (US\$/month) | |
| More than 1788 | 181 (9.8%) |
| 767-1787 | 452 (24.5%) |
| 256-766 | 644 (34.9%) |
| Less than 255 | 228 (12.4%) |
| Missing/prefer not to answer | 341 (18.5%) |
| Household income during COVID-19 pandemic |  |
| Still receiving income | 432 (23.4%) |
| Income decreased slightly | 542 (29.4%) |
| Income decreased considerably or ceased | 748 (40.5%) |
| Missing/prefer not to answer | 124 (6.7%) |
| Duration of savings to address the COVID-19 pandemic (grouped in days) |  |
| Still have savings | 1047 (56.7%) |
| It ran out between 22 to 35 days | 296 (16.0%) |
| It ran out in less than 21 days | 175 (9.5%) |
| Missing/prefer not to answer | 328 (17.8%) |
| **Factors related to the COVID-19 pandemic** |  |
| People with relatives with probable cases of SARS-CoV2 |  |
| Without probable cases of SARS-CoV2 | 1529 (82.8%) |
| With probable cases of SARS-CoV2 | 314 (17.0%) |
| Missing/prefer not to answer | 3 (0.2%) |
| The main situation during the COVID-19 pandemic |  |
| Staying at home without working | 555 (30.1%) |
| Usually does not work | 494 (26.8%) |
| Working in this period | 704 (38.1%) |
| Missing/prefer not to answer | 93 (5.0%) |
| Waiting time to purchase of most food |  |
| It is slower, takes longer to do the shopping | 1321 (71.6%) |
| It is the same as the pre-pandemic time | 210 (11.4%) |
| It is faster. It takes less time | 78 (4.2%) |
| Missing/prefer not to answer | 237 (12.8%) |
| Change in the usual place of purchase of most food |  |
| Changed the place of purchase | 875 (47.4%) |
| Did not change the place of purchase | 736 (39.9%) |
| Missing/prefer not to answer | 235 (12.7%) |
| Perception of food price of processed foods during COVID-19 pandemic |  |
| Lowered or maintained | 564 (30.6%) |
| Price increased | 1026 (55.6%) |
| Missing/prefer not to answer | 256 (13.9%) |
| **Social support in the COVID-19 pandemic** |  |
| Received some support during the COVID-19 pandemic |  |
| Yes | 432 (23.4%) |
| No | 1326 (71.8%) |
| Missing/prefer not to answer | 88 (4.8%) |
| **Perception of body condition** |  |
| Perception of weight loss during the COVID-19 pandemic |  |
| Maintained or gained weight | 1159 (62.8%) |
| Lost weight | 459 (24.9%) |
| Missing/prefer not to answer | 228 (12.4%) |
| **Food patterns and food insecurity** |  |
| Perception of consumption of minimally processed foods during the COVID-19 pandemic |  |
| Ate the same or more | 1089 (59.0%) |
| Ate less | 553 (30.0%) |
| Missing/prefer not to answer | 204 (11.1%) |
| Moderate-Severe Food insecurity (MSFI) |  |
| No | 1418 (76.8%) |
| Yes | 428 (23.2%) |

In bivariate analysis between MSFI and principal predictor variables, most were found to be significant, except for participant sex and age (table 4). The factors associated with MSFI are shown in Table 5. Among sociodemographic factors, participants who were heads of households had a prevalence of MSFI up to 1·2 times higher compared to participants who were not (aPR 1·20, CI95%: 1·00-1·44). In relation to economic factors, people whose households had an average monthly income of < 255 US$/month and between 256 to 766 US$/month in the pre-pandemic period had a prevalence of MSFI up to 3·8 (aPR 3·77 CI95%: 1·98-7·16) and 2·8 (aPR 2·80 CI95%: 1·50-5·23) times higher, respectively compared to households with an average monthly income of more than 1788 US$/month. People whose savings to address the emergency ran out in between 22 to 35 days and less than 21 days since the start of the stay-at-home order, had a prevalence of MSFI up to 1·84 (aPR 1·84, CI95%: 1·46-2·31) and 1·86 (aPR 1·86, CI95%: 1·43-2·42) times higher, respectively, compared to people with savings left to address the emergency.

**Table 4.** Bivariate analysis of potential factors associated with moderate or severe food insecurity.

| Variables | Moderate or Severe Food Insecurity |  | P* |
| --- | --- | --- | --- |
|  | No (total=1418)<br>n (%) | Yes (total=428)<br>n (%) |  |
| **Sociodemographic factors** |  |  |  |
| Head of household |  |  | 0.018 |
| No | 914 (78.3%) | 253 (21.7%) |  |
| Yes | 463 (73.4%) | 168 (26.6%) |  |
| **Economic factors** |  |  |  |
| Household income pre-pandemic period (US\$/month) | | | < 0.001 |
| More than 1788 | 168 (92.8%) | 13 (7.2%) |  |
| 767-1787 | 403 (89.2%) | 49 (10.8%) |  |
| 256-766 | 461 (71.6%) | 183 (28.4%) |  |
| Less than 255 | 113 (49.6%) | 115 (50.4%) |  |
| Household income during COVID-19 pandemic |  |  | < 0.001 |
| Still receiving income | 395 (91.4%) | 37 (8.6%) |  |
| Income decreased slightly | 457 (84.3%) | 85 (15.7%) |  |
| Income decreased considerably or ceased | 466 (62.3%) | 282 (37.7%) |  |
| Duration of savings to address the COVID-19 pandemic (grouped in days) |  |  | < 0.001 |
| Still have savings | 908 (86.7%) | 139 (13.3%) |  |
| It ran out between 22 to 35 days | 161 (54.4%) | 135 (45.6%) |  |
| It ran out in less than 21 days | 85 (48.6%) | 90 (51.4%) |  |
| **Factors related to the COVID-19 pandemic** |  |  |  |
| People with relatives with probable cases of SARS-CoV2 |  |  | < 0.001 |
| Without probable cases of SARS-CoV2 | 1200 (78.5%) | 329 (21.5%) |  |
| With probable cases of SARS-CoV2 | 216 (68.6%) | 98 (31.2%) |  |
| The main situation during the Covid-19 pandemic |  |  | < 0.001 |
| Staying at home without working | 385 (69.4%) | 170 (30.6%) |  |
| Usually does not work | 384 (77.7%) | 110 (22.3%) |  |
| Working in this period | 581 (82.5%) | 123 (17.5%) |  |
| Waiting time to purchase of most food |  |  | 0.017 |
| It is slower, takes longer to do the shopping | 1000 (75.7%) | 321 (24.3%) |  |
| It is the same as the pre-pandemic time | 173 (82.4%) | 37 (17.6%) |  |
| It is faster. It takes less time | 67 (85.9%) | 11 (14.1%) |  |
| Change in the usual place of purchase of most food |  |  | 0.013 |
| Changed the place of purchase | 656 (75.0%) | 219 (25.0%) |  |
| Did not change the place of purchase | 590 (80.2%) | 146 (19.8%) |  |
| Perception of food price of processed foods during the COVID-19 pandemic |  |  |  |
| Lowered or maintained | 465 (82.5%) | 99 (17.6%) | < 0.001 |
| Price increased | 757 (73.8%) | 269 (26.2%) |  |
| **Social support in the COVID-19 pandemic** |  |  |  |
| Received some support during the COVID-19 pandemic |  |  | < 0.001 |
| Yes | 284 (65.7%) | 148 (34.3%) |  |
| No | 1066 (80.4%) | 260 (19.6%) |  |
| **Perception of body condition** |  |  |  |
| Perception of weight loss during the COVID-19 pandemic |  |  | < 0.001 |
| Maintained or gained weight | 929 (80.2%) | 230 (19.8%) |  |
| Lost weight | 318 (69.3%) | 141 (30.7%) |  |
| **Food patterns and food insecurity** |  |  |  |
| Perception of consumption of minimally processed foods during the COVID-19 pandemic |  |  | < 0.001 |
| Ate the same or more | 944 (86.7%) | 145 (13.3%) |  |
| Ate less | 323 (58.4%) | 230 (41.6%) |  |

\* Chi-square test.

**Table 5.**
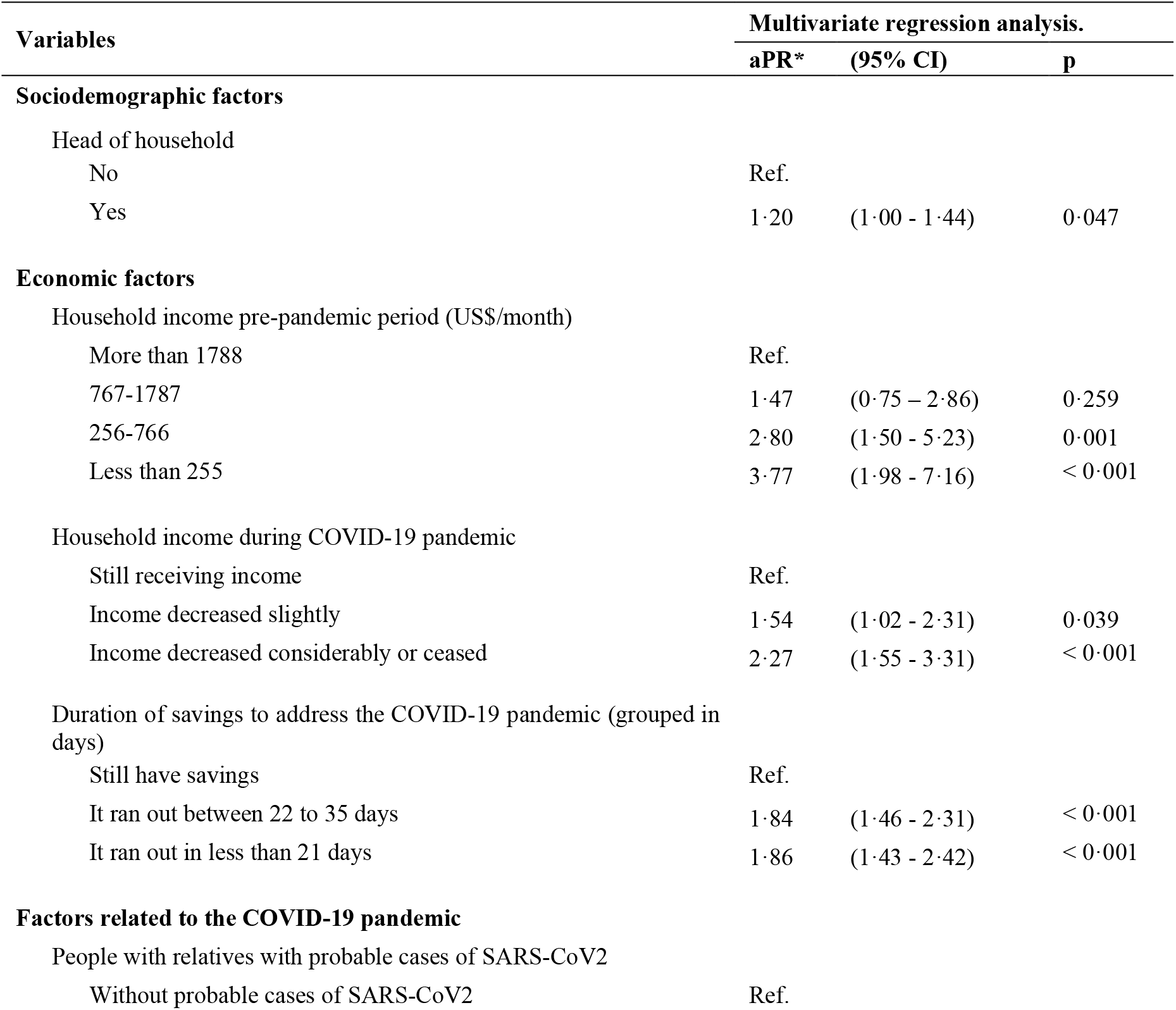
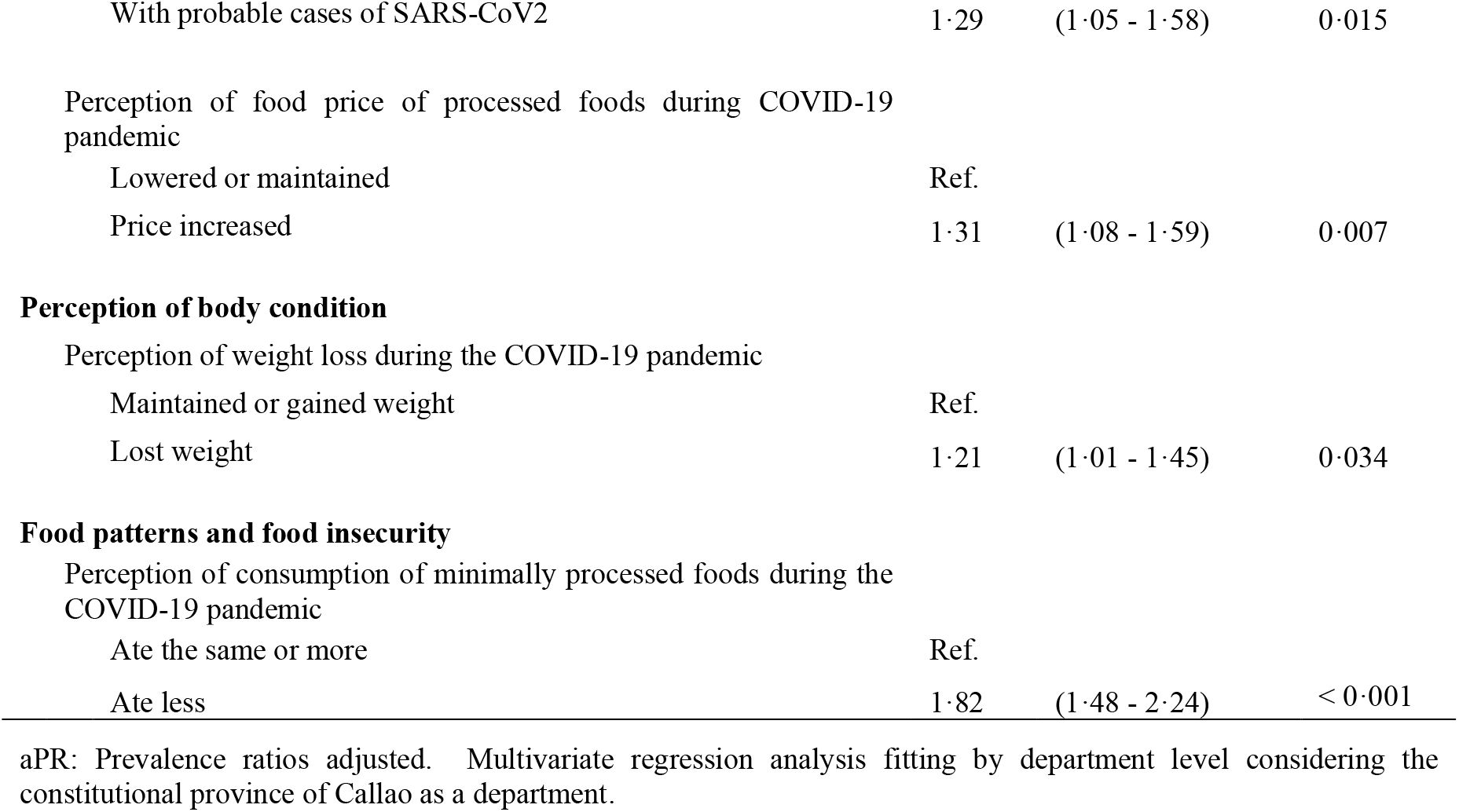
Prevalence ratios of factors associated with moderate-severe food insecurity. Peru

On the other hand, participants with relatives who were probable cases of SARS-CoV2 had a prevalence of MSFI up to 1·29 (aPR 1·29, CI95%: 1·05-1·58) times higher relative to those who did not. Participants who reported a loss of weight during the pandemic period had a prevalence of MSFI 1·21 (aPR 1·21, CI95%: 1·01-1·45) times higher than those who maintained or gained weight. Lastly, participants who reported consuming less natural or minimally processed food had a prevalence of MSFI 1·82 (aPR 1·82, CI95%: 1·48-2·24) times higher than those who ate the same or more of that food group. Participants who reported increases in processed foods prices had a prevalence of MSFI 1.31 (aPR 1·31; 95%CI, 1·08-1·59) times higher than those who reported that prices were maintained or decreased.

## DISCUSSION

The present study found a prevalence of MSFI of 23·2%. Factors associated with it were having an income below the minimum living wage in the pre-pandemic period, having experienced a substantial reduction in income during the pandemic, ran out of savings in the first 21 days of the pandemic period, being a head of household, having relatives who were probable cases of SARS-CoV2, perceiving weight loss during pandemic period, reporting that they ate less natural or minimally processed food, and having perceived an increase in prices of processed foods.

In Peru in 2014, the reported prevalence of MSFI was 29·9%;^(35)^ however, our study found a prevalence of MSFI of 23·2% in the first 30-45 days of stay-at-home order. Although these results are not comparable with MSFI initial values, the prevalence of MSFI in this study may be underestimated because of some particular characteristics of the population analysed. For example, 73·3% had a university education, only 12·4% reported having monthly income less than 255 US$/month before the pandemic, and participants required internet access and a social network account to access the survey, attributes that low-income people generally do not have and they are precisely the population most at risk of suffering food insecurity.^(22, 26)^ Other studies that have evaluated Food Insecurity in the context of the COVID-19 pandemic using FIES have reported higher MSFI prevalence. In Jordan, the prevalence of MSFI was 59·2%,^(36)^ in Kenya and Uganda MSFI increased from 50% to 88% and from 43% to 87%, from before to during the pandemic, respectively.^(15)^ Our study measures individual MSFI; however, there is already evidence of the effect stay-at-home order on household MSFI. In Mexico, it was reported that the prevalence of household MSFI increased from 24·2% in 2018 to 30·2% in June 2020.^(31)^ Vermont also reported is an increase of MSFI from 18·3% pre-pandemic to 24·4% during the pandemic.^(14)^ In rural Bangladesh the prevalence of MSFI increased from 8·3% pre-pandemic to 51·8% during the pandemic.^(11)^

Our study is one of the first investigations into MSFI during the stay-at-home order due to COVID-19 pandemic in Peru. We found that up to 40·5% of participants experienced a considerable reduction in their income, similar results were reported by the Young Lives study in Peru with 37% of households affected by a salary cut or suspension without payment.^(8)^ These high percentages of income losses could be related to the fact that in Peru, 50% of income came from informal employment in the pre-pandemic period.^(5)^ This could explain our results, i.e., those who had lower income (compared to high-income people) before the pandemic, had less capacity to save money to face emergency situations, and had substantial income reduction during the pandemic, are shown to be more likely to have MSFI. These findings are consistent with other studies, for example, in Jordan it was reported that people who had a monthly income per capita below the poverty line were more likely to have moderate (OR: 5·33; 95% CI: 4·44–6·40) and severe food insecurity and severe food insecurity (OR: 6·87; 95% CI: 5·542–8·512).^(36)^ Similarly, other studies that explored determinants of food insecurity during the COVID pandemic reported a higher risk of MSFI in low-income people,^(37)^ people who lost their job,^(14, 38)^ or people with substantial income loss.^(39)^

The Peruvian government implemented social aid programs aimed at poor populations to help them face the stay-at-home order; this aid consisted mainly of direct payment of 760 Peruvian soles (208·5 US$), early withdrawal from the private retirement pension, and food donations. In our study, we found that most respondents reported not having been part of this beneficiary population. For example, only 15·7% received some type of support from the government and of these only 25·6% was allocated to people with pre-pandemic monthly income below the minimum living wage (less than 255 US$) and 55·6% to people with monthly income from 256 to766 US$ (data not shown in tables). We do not have information on the percentage of the vulnerable population that did not receive government aid to face the stay-at-home order in Peru. However, it has been reported that the data source used to identify vulnerable households was designed for regular contexts and does not allow for the identification of families that abruptly entered poverty due to the mandatory stay-at-home order.^(5)^ Problems with receiving money from social assistance were also reported, because nearly 14 million adult Peruvians (59·8% of the total adult population) did not have a bank account.^(40)^ For this reason, our results invite further analysis and discussion on the targeting and delivery of government social aid to vulnerable populations during the pandemic.

On the other hand, the high food price perception during the pandemic (55·6% participants reported an increase in the price of processed food in our study) may have limited the capacity to buy or access food during the pandemic. Loss of jobs^(11, 41)^ and increase in food prices^(12)^ are some of the side effects of stay-at-home orders reported by other studies.

In this study we found that people who had reported consuming less minimally processed food were more likely to experience MSFI, this may be due to reduced access to food in terms of quantity and quality, increased food price or having some fear of SARS-CoV2 infection in markets because products are not packaged.^(42)^

It is possible that one of the reasons heads of households were more likely to suffer from MSFI could be stress and anxiety related to getting food and having to prioritize feeding their family over themselves. ^(7)^ This result could be related to the perception of losing weight that, in our study, was associated with MSFI.

Our findings demonstrate a greater chance of experiencing MSFI in participants with relatives who were probable cases of SARS-CoV2. Even though there is no evidence about a direct relationship between MSFI and SARS-CoV2, we suggest that there could be an indirect relationship, as household economic resources could be allocated primarily to treatment or care of the infected household member instead of providing food at home.

The study has some limitations. Because it is a cross-sectional study; it cannot establish causal relationships. There might be bias as participants were not randomly selected, thus the results shown are only generalizable to our study population.

It is important to discuss how the sample was obtained. This study employed a web-based survey where participants were not randomly selected. Ideally, we expected to have a sample with sociodemographic characteristics similar to the Peruvian national population (21·2% and 20·8% within the poorest and poor quintiles, respectively; 28·8% residing in the province of Metropolitan Lima; 60% of women between 15-49 years old having at least secondary education and around 40% having higher education).^(43, 44)^ However, our sample consisted mainly of women (74·9%) and people who residing in the province of Metropolitan Lima (67·7%); with only 12·9% of those women have at least secondary level education, 87·1% have higher or university education (data not shown) and with 12·4% of households reporting an average monthly income of less than 255 US$/month in pre-pandemic period (similar to the poorest quintile of the Peruvian national population). Considering these discrepancies between the general Peruvian population and our study sample, we can mention that this study’s sample has a similar distribution to Peruvian population that live in Metropolitan Lima, with an upper-middle income socioeconomic status and it is not generalizable to the whole Peru. However, we showed important evidence on MSFI in the context of the stay-at-home order due to the COVID-19 pandemic in Peru.

In conclusion, people who went through MSFI were economically vulnerable both before and during stay-at-home order due to COVID-19 pandemic in Peru. Factors most strongly associated with MSFI included being a head of household, consuming less minimally processed food, and perceiving losing weight. It is necessary to implement effective social assistance policies to prevent or mitigate these adverse effects.

## Data Availability

Requests of survey data will be considered for research purposes, while does not conflict with other requests by the study authors. Requests should be made to the correspondent author.

## AUTHORSHIP CONTRIBUTIONS

OCH, MP and JLC led the design of the study. JLC, OCH, SH, GD, AA, and JPG contributed to designing and validating the instrument and collecting of data. JLC and OAE carried out the data analysis. DAA, JPA and MP were responsible for the interpretation of the results. JLC contributed to the initial drafting of the manuscript based on comments from DAA, MP, and JPA. All authors contributed to the interpretation of data, critical revision of the manuscript and approved the final version.

## DATA SHARING

Requests of survey data will be considered for research purposes, while does not conflict with other requests by the study authors. Requests should be made to Jorge L. Cañari-Casaño.

## ACKNOWLEDGMENTS

We want to thank Rikolto in Latin America and the Belgian cooperation for their contribution in designing the advertising campaign on social media. We also thank Colegio de Nutricionistas del Perú, Universidad Nacional Mayor de San Marcos and all the public Peruvians universities that sharing at not cost our survey online from their official social media pages.

JLC is a doctoral student studying an Epidemiological Research Doctorate at Universidad Peruana Cayetano Heredia under FONDECYT/CIENCIACTIVA scholarship EF033-235-2015 and supported by training grant D43 TW007393 awarded by the Fogarty International Center of the US National Institutes of Health.

We also express our acknowledgement to Hilary Creed for her comments on our protocol of study and John Nesemann, for his review of the English version of our manuscript.

